# A clinical study to evaluate the safe and effective use of Stethoglove, a single use stethoscope cover to reduce pathogen transmission during auscultation

**DOI:** 10.1101/2023.04.07.23288292

**Authors:** Timo Z. Nazari-Shafti, Heike Meyborg, Jasper Iske, Maximilian Schloss, Fabian Seeber, Aljona Friedrich, Vasileios Exarchos, Anja Richter, Volkmar Falk, Maximilian Y Emmert

**Affiliations:** Deutsches Herzzentrum der Charité (DHZC), Department of Cardiothoracic and Vascular Surgery, Berlin, Germany; Charité – Universitätsmedizin Berlin, corporate member of Freie Universität Berlin and Humboldt-Universität zu Berlin, and Berlin Institute of Health, Berlin, Germany; German Centre for Cardiovascular Research (DZHK), Partner Site Berlin, Berlin, Germany

**Keywords:** stethoscope, nosocomial infections, auscultation, transmission, polymer

## Abstract

**Objectives:** Stethoscopes carry a significant risk for pathogen transmission. Here, the safe use and performance of Stethoglove^®^, a non-sterile, single-use cover for stethoscopes, that is impermeable for pathogens, was investigated by different healthcare professionals (HCPs) in the postoperative care setting of an intensive care unit (ICU).

**Methods:** Fifty-four patients underwent routine auscultations with the use of Stethoglove^®^. The participating HCPs (n=34) rated each auscultation with Stethoglove^®^ on a 5-point Likert scale. The mean ratings of acoustic quality and Stethoglove^®^ handling were defined as primary and secondary performance endpoint.

**Results:** 534 auscultations with Stethoglove^®^ were performed (average 15.7/user) on the lungs (36.1%), the abdomen (33.2%), the heart (28.8%) or other body-sites (1.9%). No adverse device-effects occurred. The acoustic quality was rated at 4.2±0.7 (mean) with a total of 86.1% of all auscultations being rated at least as 4/5, and with no rating as below 2. The Stethoglove^®^ handling was rated at 3.7±0.8 (mean) with a total of 96.4% of all auscultations being rated at least 3/5.

**Conclusions:** Using a real-world setting, this study demonstrates that Stethoglove^®^ can be safely and effectively used as cover for stethoscopes during auscultation. Stethoglove^®^ may therefore represent a useful and easy-to-implement tool for preventing stethoscope-mediated infections.

## INTRODUCTION

Healthcare-associated infections (HAIs) represent a significant risk for patients ^1^. In the United States, the Center for Disease Control (CDC) reports that about 1.7 million patients are estimated to be affected by HAIs every year while they are undergoing in-hospital treatment for other health problems, and about 100,000 patients (approx. 6%) are dying from such HAIs ^2^. Widely regarded as the “physician’s third hand” the stethoscope plays a critical role in the transmission of pathogens ^3-8^, and importantly, they harbor similar contamination levels as the HCP’s hand ^2,6,9^. Notably, with more than 5 billion auscultations per year in the US alone, it represents the most often used medical device in daily clinical practice ^2^.

Stethoscopes are crucial for both, in hospital and outpatient environments. They are not only used for standard physical assessments, but also for the examination of special medical conditions such as the diagnosis of abdominal or heart diseases or during routine postoperative care, e.g., in intensive care units (ICU).

Disinfection of stethoscopes with isopropyl alcohol is recommended and may be effective at eliminating many pathogens ^3,5,10^. However, even though stethoscope cleaning guidelines and recommendations from the CDC are available (i.e. continuous wiping with isopropyl alcohol for at least 60 seconds), the overall compliance of health care professionals (HCPs) in the daily clinical routine is reported to be extremely low ^4,11,12^.

A recent study from Boulee and colleagues found that stethoscopes are most often not cleaned between patient auscultations. Moreover, in cases where cleaning was performed it only complied in 4% of the cases with the current CDC guidelines ^11,13^. The reasons for this non-compliance are multifactorial (e.g. lack of time, material and reminders) ^12^. To date, many initiatives to improve stethoscope hygiene have widely failed, and commonly accepted “best practices” are yet to be developed. In addition, even if current stethoscope cleaning guidelines would be practiced, highly virulent pathogens such as methicillin-resistant *S. aureus* (MRSA) or *C. difficile* spores are likely to persist on the stethoscope diaphragm thereby posing a significant risk to the next patient ^14^.

Alternative strategies to enhance stethoscope hygiene have been limited so far ^15^. However, driven by the ongoing COVID-19 pandemic, the search for new stethoscope protection technologies and concepts has recently regained reasonable attention ^9^. One promising approach are microbiological barriers (e.g. single-use, disposable covers) shielding the stethoscope diaphragm direct patient contact ^15,16^. However, while such concepts may indeed have the potential to improve stethoscope hygiene, they should not alter auscultation accuracy by impairing the acoustic quality of auscultation sounds. Available data is scarce in this regard ^16^. Another important requirement for such a device is its ease of use as to enable simple and rapid implementation into the clinical routine workflow ^2^.

Recently, the medical device Stethoglove^®^, a non-sterile, single-use hygienic cover for stethoscopes has been developed. It is impermeable for microorganisms thereby preventing stethoscope-mediated patient-to-patient transmission of pathogens and preserves the acoustic quality of auscultations. Stethoglove^®^ can be easily pulled over the stethoscope’s bell, part of the tube(s), while being held in the hand of the user to also shield the operating hand during auscultation.

In the present clinical study, the safe use and performance of Stethoglove^®^ was investigated by different HCPs under real-life conditions in the daily routine setting of an ICU for cardiac surgery patients.

## PATIENTS AND METHODS

### Study design and objectives

The aim of this single-arm, open-label clinical study was to investigate the safe use and performance of Stethoglove^®^ by HCPs under real-life conditions in an ICU for cardiovascular surgery patients. The objectives of the study were to assess 1.) the acoustic quality of auscultations with Stethoglove^®^ (primary performance endpoint); and 2.) the overall usability of Stethoglove^®^ (secondary performance endpoint) in the daily clinical routine by different HCPs (user groups) including physicians and nurses which were further characterized by their level of clinical experience. The study was approved by the local ethics committee (21-702-MP Mono -IVE17) and the federal authority (94.1-03-5660-13526).

### Patients and study course

Upon hospital admission on the day prior to their elective cardiovascular surgery adult patients (≥ 18 years) were asked for informed consent to participate in the study. Patients who provided informed consent were screened for inclusion and exclusion criteria (see definitions in supplementary file). In included patients, the participating users performed auscultations as part of their standard postoperative care with Stethoglove^®^.

After each auscultation the users entered the following information and rating on a study-specific and access-restricted page embedded into the patients’ electronic medical record:

1. Auscultated organ or body site
  1. Heart
  2. Lung
  3. Abdomen
  4. Other
2. Rating of the acoustic quality of the auscultation with Stethoglove^®^ (on a 5-point Likert scale)

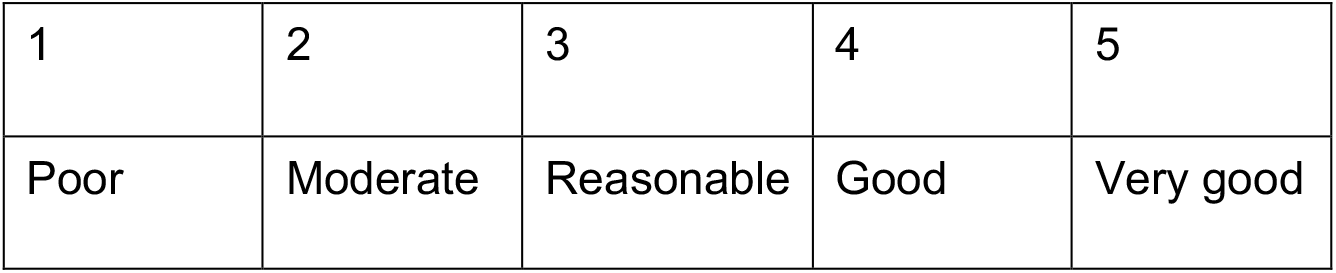
3. Rating of the Stethoglove^®^ handling as part of the routine working practice (on a 5-point Likert scale)

|  |  |  |  |  |
| --- | --- | --- | --- | --- |
| 1 | 2 | 3 | 4 | 5 |
| Poor | Moderate | Reasonable | Good | Very good |

3. Rating of the Stethoglove<sup>®</sup> handling as part of the routine working practice (on a 5-point Likert scale)
|  |  |  |  |  |
| --- | --- | --- | --- | --- |
| 1 | 2 | 3 | 4 | 5 |
| Poor | Moderate | Reasonable | Good | Very good |

### Users

A total of 34 physicians and nurses with different levels of professional experience participated in this study and were assigned to one of the following groups:

<u>User group 1:</u> Fully trained physicians (e.g., senior physician, board-certified physician, medical specialist, physicians with at least 8 years of working experience) (n=8)

<u>User group 2:</u> Physicians in specialty training (e.g., residents) (n=7)

<u>User group 3:</u> Fully trained nurses with at least 10 years of working experience (n=10)

<u>User group 4:</u> Fully trained nurses with less than 10 years of working experience (n=9)

### Device Description

Stethoglove^®^ (Stethoglove GmbH, Hamburg, Germany) is a hygienic stethoscope cover and a safe and effective solution to minimize the risk of patient-to-patient infections caused by contaminated stethoscopes in a health care environment (Figure 1, product images).

**Figure 1:**
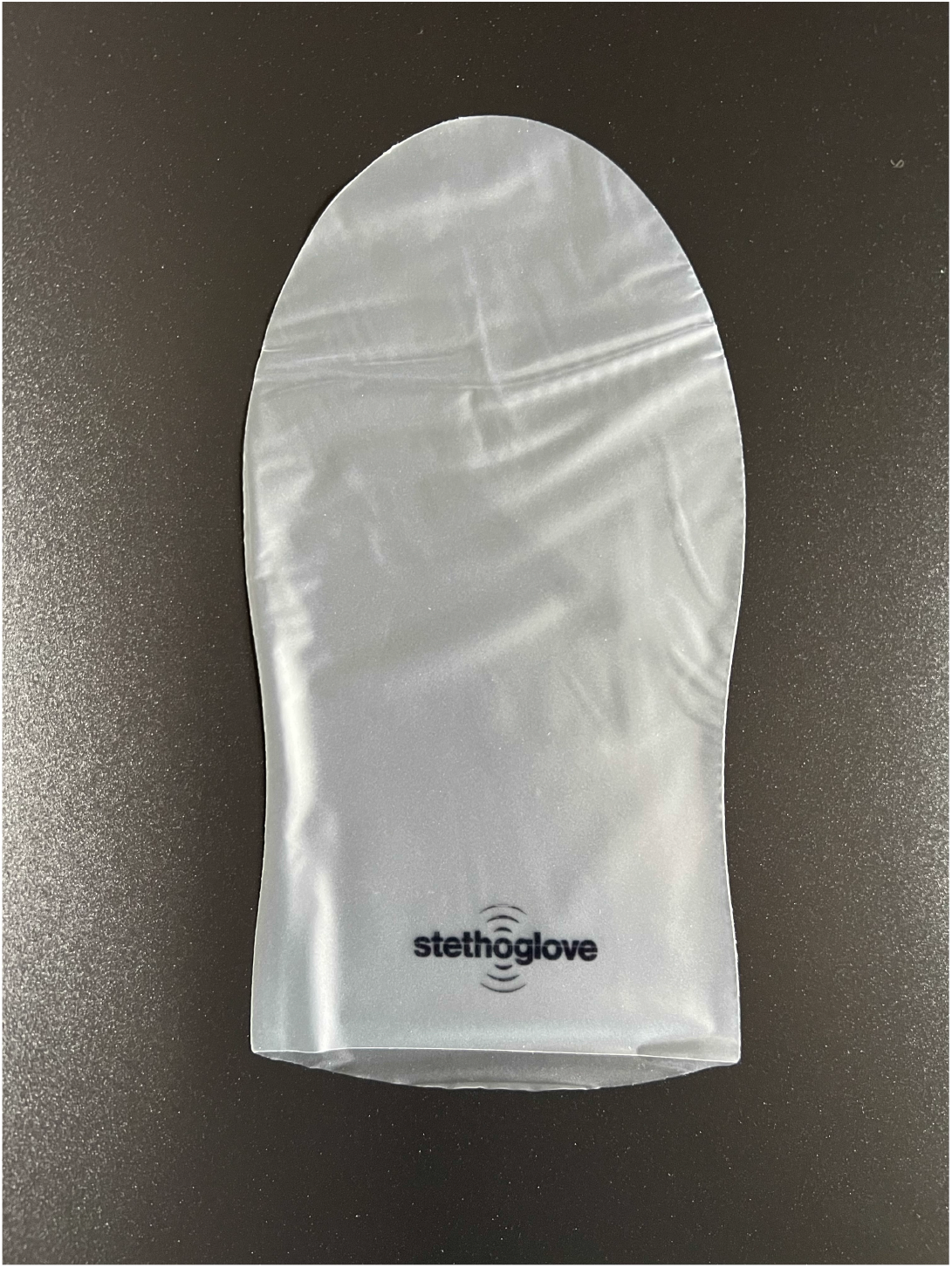
Product image of single use glove.

The non-sterile, single-use glove is pulled over the stethoscope’s bell and part of the tube(s), while held in the hand of the user (Figure 2, product handling images).

**Figure 2:**
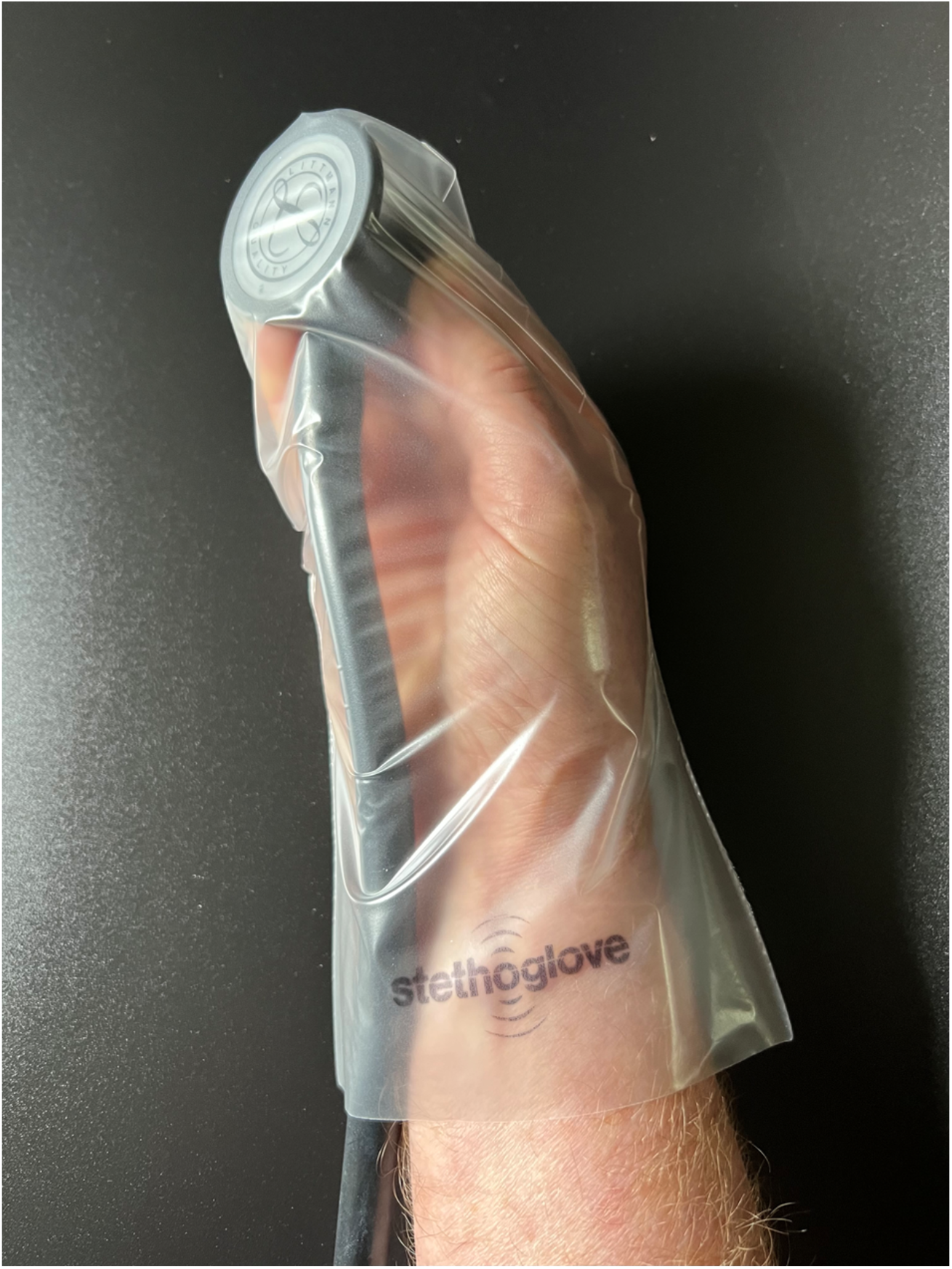
Product handling images.

Stethoglove^®^ is made of thermoplastic urethane (TPU) and is latex- and powder-free. The TPU material is impermeable for microorganisms such as bacteria, fungi and viruses. Such impermeability was demonstrated with decontaminated stethoscope bells wrapped in a Stethoglove^®^ cover, which were exposed for 30 minutes to confluently grown bacterial (*S. aureus, B. diminuta*) and fungal (*A. niger, C. albicans*) cultures on agar plates. Swabbing of the unwrapped stethoscope diaphragm post exposure revealed no contaminations [unpublished data]. Viral impermeability was tested in accordance with Method C of ISO Norm 16604 (2004) using the blood-borne pathogen Bacteriophage Phi-X1274. Cut-outs from four different areas of a Stethoglove^®^ cover were exposed to the virus-containing suspension in a penetration cell and a sequence of pressures ranging from 0 to 20 kpa was applied each for 5 minutes. No virus penetration was detected [unpublished data].

Regarding acoustic quality Stethoglove^®^ has demonstrated excellent transmission of auscultation sounds in both lab tests and a usability test with HCPs [unpublished data]. By combining this impermeability for microorganism with the preserved acoustic quality of auscultation sounds Stethoglove^®^ ensures proper and safe patient-to-patient use of stethoscopes by HCPs in their day-to-day practice and thus improves patient protection from HAIs.

### Study endpoints

In this study the following performance and safety endpoints were defined:

- The primary performance endpoint was the rating of the acoustic quality of auscultations with Stethoglove^®^ by all users
- The secondary performance endpoint was the rating of the handling of Stethoglove^®^ as part of the routine working practice by all users
- The safety endpoint was the frequency of observed adverse device effects or serious adverse device effects

In addition, the influence of gender, BMI and auscultated organ/body site on the primary and secondary performance endpoints were evaluated

### Sample size calculation and statistical analysis

The continuous parameter “rating of the acoustic quality during auscultation measured on a 5-point Likert scale (poor, moderate, reasonable, good, very good)” was used as primary performance endpoint. No verification of a pre-defined hypotheses was planned. Instead, a total of 508 recorded auscultations with Stethoglove^®^ was calculated to be required based on the following assumptions: a sampling error of 5% (precision), a prognostic accuracy of 95% (reliability), an expected mean of 4 (assessment of acoustic quality, “good”) and a variance of 2.3 points (homogeneity).

Both the primary and secondary performance endpoints were first evaluated descriptively and then stratified by the following influencing factors: patient gender, BMI and auscultated organ/body site. All analyses were performed within all users, and within each user group. Missing values were not replaced.

All measured differences between groups were investigated for numeric variables by means of a two-sided t-Test for independent samples (comparison of 2 groups) and a one-factorial ANOVA (Welch-Test, comparison of more than 2 groups) with a type I error of 0.05. Since no formal hypothesis testing was done, all p-values were interpreted in a descriptive manner.

## RESULTS

### Baseline characteristics, demographics and safety

A total of 54 patients were enrolled in this study and 534 auscultations with Stethoglove^®^ were performed with an average of 9.9 auscultations per patient. Patient characteristics are shown in Table 1.

**Table 1.** Patient demographics.

| Variable |  | Total Patients<br>(N=54) | Percentage<br>(%) | Statistics |
| --- | --- | --- | --- | --- |
| Gender | Female | 15 | 27.8 |  |
|  | Male | 39 | 72.2 |  |
| Body Mass<br>Index | N |  |  | 54 |
|  | Mean ±SD |  |  | 27.8 ± 5.2 |
|  | Median |  |  | 27.4 |
|  | Range (Min – Max) |  |  | 19.8 - 43.8 |
|  | Normal weight (18.5 - 24.9) | 19 | 35.2 |  |
|  | Overweight (25.0 - 29.9) | 19 | 35.2 |  |
|  | Class I obesity (overweight) (30.0 - 34.9) | 13 | 24.1 |  |
|  | Class II obesity (obesity) (35.0 - 39.9) | 0 | 0.0 |  |
|  | Class III obesity (extreme obesity) (≥40.0) | 3 | 5.6 |  |

1 \*SD= Standard Deviation

More males (n=39) than females (n=15) participated in this investigation which represents the typical gender distribution in cardiovascular patients. The mean BMI in the patient cohort was

27.8 ±5.2 kg/cm^2^. The majority of patients (n=35) were overweight, 13 patients suffered from class I and 3 patients from class III obesity.

The rate of auscultations per organ and body site, were evenly distributed between lung (36.1%), abdomen (33.2%) and heart (28.8%), while only 1.9% of the auscultations were performed at other body sites (e.g., carotid artery) (Table 2). No adverse device effects (ADEs) were reported during the study.

**Table 2:** Distribution of auscultations with Stethoglove^®^ by body site and user group.

| Variable |  | Total auscultations (N=534) | Percentage (%) |  |
| --- | --- | --- | --- | --- |
| Organ/Body site | Abdomen | 177 | 33.2 |  |
|  | Heart | 154 | 28.8 |  |
|  | Lung | 193 | 36.1 |  |
|  | Other | 10 | 1.9 |  |
|  |  |  |  | Average no. per user |
| User Group | No. 1: Fully trained physicians (N=8) | 137 | 25.7 | 17.1 |
|  | No. 2: Physicians in speciality training (N=7) | 114 | 21.4 | 16.3 |
|  | No. 3: Fully trained nurses with ≥10 years of experience (N=10) | 152 | 28.5 | 13.8 |
|  | No. 4: Fully trained nurses with <10 years of experience (N=9) | 131 | 24.5 | 16.4 |
|  | **All (N=34)** | **534** | **100** | **15.7** |

### Acoustic quality of auscultations with Stethoglove^®^

#### Primary Performance Endpoint

The mean rating of the acoustic quality of auscultations with Stethoglove^®^ by all users was 4.2 ± 0.7 (Median 4.0; range 2-5) on a 5-point Likert scale (Table 3). A total of 86.1% of all auscultations with Stethoglove^®^ were rated at least “good” (score of 4 or higher) with comparable results across user groups ranging from 88.3% for fully trained physicians (group 1), 85.1% for physicians in speciality training (group 2), 87.5% for nurses with 10 or more years of professional experience (group 3), and 83.1% for nurses with less than of professional experience (group 4), respectively (Table 3, Figure 3). No auscultation was rated as “poor” (score of 1). For two out of 534 auscultations no rating was recorded.

**Table 3:** Percentage of Stethoglove^®^ handling rated as “reasonable”, “good” and “very good”.

| User group | Ratings of handling (% and n) |  |  |  |  |
| --- | --- | --- | --- | --- | --- |
| | Mean ±SD | “Reasonable”<br>(score of 3) | “Good”<br>(score of 4) | “Very good”<br>(score of 5) | Combined |
| All users | 3.7 ±0.8<br>(n=533*) | 40.8%<br>(n=218) | 34.6%<br>(n=185) | 21.0%<br>(n=112) | 96.4%<br>(n=515) |
| User group 1 | 4.0 ±0.9<br>(n=136) | 32.9%<br>(n=45) | 24.8%<br>(n=34) | 39.4%<br>(n=54) | 97.1%<br>(n=133) |
| User group 2 | 3.7 ±0.7<br>(n=114) | 34.2%<br>(n=39) | 52.6%<br>(n=60) | 10.5%<br>(n=12) | 97.3%<br>(n=111) |
| **User group 3** | 3.8 ±0.8<br>(n=152) | 21.7%<br>(n=33) | 50.0%<br>(n=76) | 20.4%<br>(n=31) | 92.1%<br>(n=140) |
| **User group 4** | 3.3 ±0.7<br>(n=131) | 77.1%<br>(n=101) | 11.5%<br>(n=15) | 11.5%<br>(n=15) | 100%<br>(n=131) |

\* For one out of 534 auscultations (0.2%) no rating was recorded

**Figure 3:**
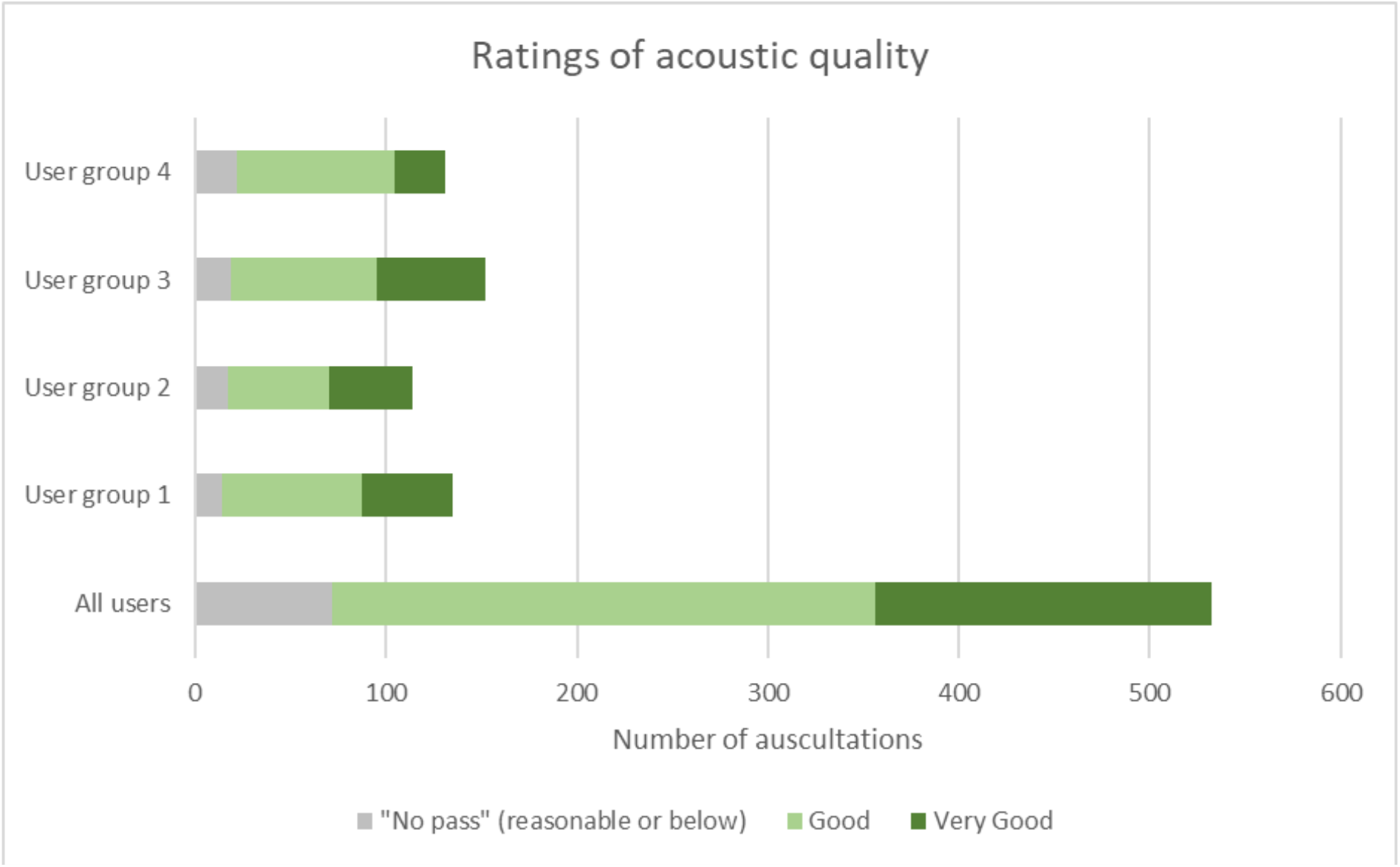
Distribution of ratings of auscultation quality among the four user groups. User group 4: Fully trained nurses with <10 years of experience, user group 3: nurses with >10 years of experience, user group 3: Physicians in specialty training (e.g., residents), user group 1: Fully trained physicians (e.g., senior physician, board-certified physician, medical specialist, physicians with at least 8 years of working experience)

#### Acoustic quality of auscultations with Stethoglove^®^ stratified by gender, BMI and auscultated organ/body site

The influence of the independent variables gender, BMI, and auscultated organ/body site on the rating of the acoustic quality of auscultations with Stethoglove^®^ by all users was evaluated separately. The rating of the acoustic quality in male and female patients was comparable with a mean score of 4.2 ± 0.7 in males and 4.0 ± 0.7 in females (Median 4.0; range 2-5 for both) (Suppl. Table S1).

When stratified by BMI the mean rating of the acoustic quality was 4.3 ± 0.6 (for normal weight patients), 4.2 ± 0.8 (for overweight patients), 4.0 ± 0.6 (for class I obesity patients) and 3.8 ± 0.8 (for class III obesity patients) (Median 4.0; range 2-5 for all BMI groups) (Suppl. Table S2). When stratified by auscultated organ/body site the mean rating of the acoustic quality was 4.3 ± 0.7 for lungs, 4.2 ± 0.7 for abdomen, and 4.1 ± 0.6 for the heart (Median 4.0; range 2-5 for all three). For “other” body sites including the carotid artery, femoral artery and stomach, a lower mean rating was assigned (3.3 ± 0.7; Median 3.0; range 2-4), however, the underlying number of auscultations was substantially lower than for the lung, abdomen and heart (Suppl. Table S3).

#### Acoustic quality of auscultations with Stethoglove^®^ by user groups

The ratings were consistent across all user groups, i.e., 4.2 ± 0.7 for user groups 1-3, and 4.0 ± 0.7 for user group 4.

### Evaluation of Stethoglove^®^ usability (handling)

#### Secondary performance endpoint

The mean rating of the handling of Stethoglove^®^ in the daily clinical routine by all users was 3.7 ± 0.8 (Median 4.0; range 2-5) (Table 4. A total of 96.4% of all auscultations was rated to be at least “reasonable” by all users, while no auscultation was rated as “poor” (score of 1) (Table 4, Figure 4). This suggests a high level of acceptability to use Stethoglove^®^ throughout all user groups. No handling was rated as “poor” (score of 1). For one out of 534 auscultations no rating of the handling was recorded.

**Table 4:** Percentage of auscultations with Stethoglove^®^ where the acoustic quality was rated as “good” and “very good”.

| User group | Ratings of acoustic quality (% and n) |  |  |  |
| --- | --- | --- | --- | --- |
| | Mean ±SD | “Good”<br>(score of 4) | “Very good”<br>(score of 5) | Combined |
| **All users** | 4.2 ±0.7 (n=532*) | 53.2%<br>(n=284) | 33.0%<br>(n=176) | 86.1%<br>(n=460) |
| **User group 1** | 4.2 ±0.7 (n=135) | 53.3%<br>(n=73) | 35.0%<br>(n=48) | 88.3%<br>(n=121) |
| **User group 2** | 4.2 ±0.7 (n=114) | 46.5%<br>(n=53) | 38.6%<br>(n=44) | 85.1%<br>(n=97) |
| **User group 3** | 4.2 ±0.7 (n=152) | 50.0%<br>(n=76) | 37.5%<br>(n=57) | 87.5%<br>(n=133) |
| **User group 4** | 4.0 ±0.7 (n=131) | 62.6%<br>(n=82) | 20.6%<br>(n=27) | 83.2%<br>(n=109) |

\* For two out of 534 auscultations (0.3%) no rating was recorded

**Figure 4:**
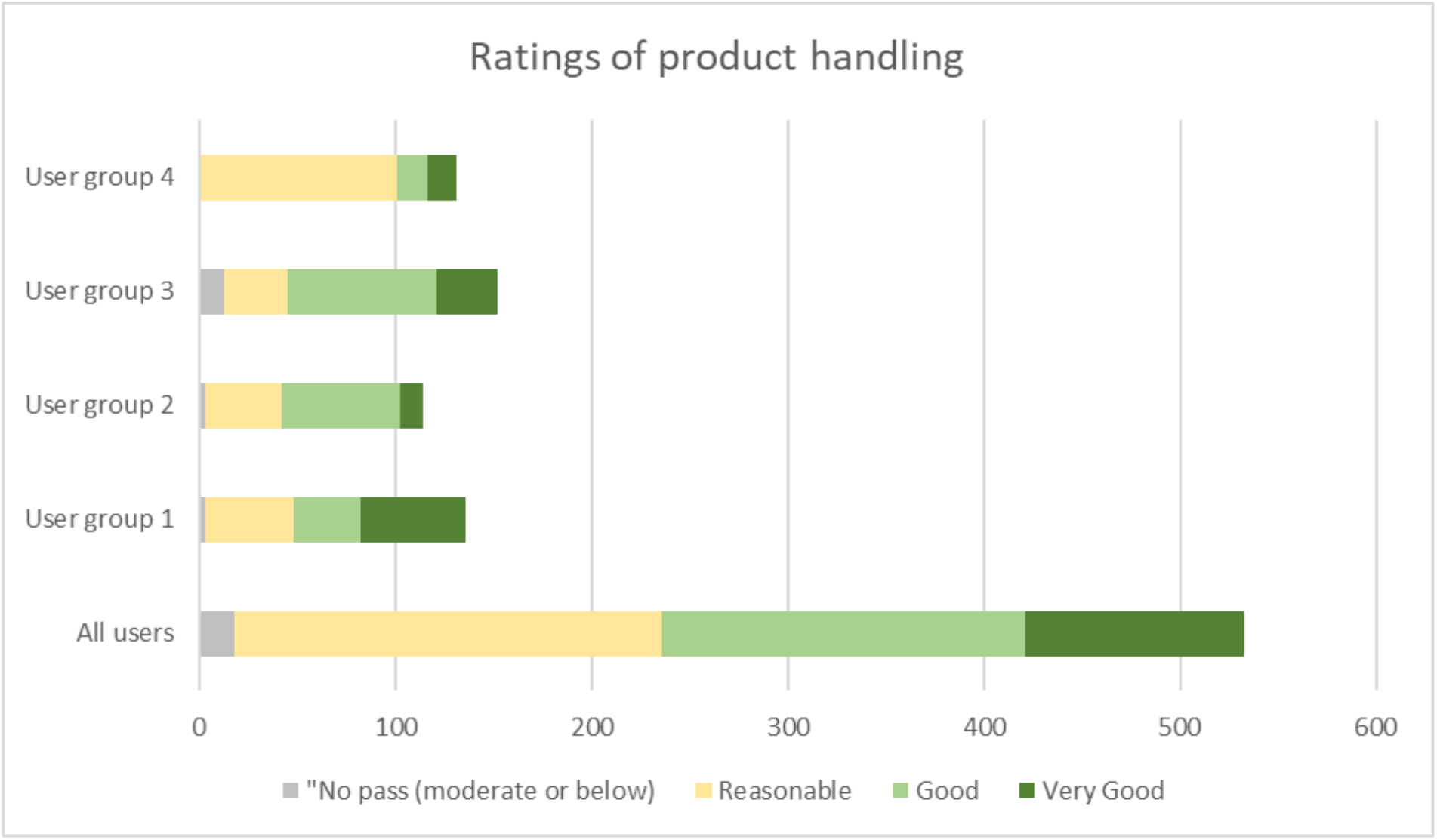
Distribution of ratings of product handling among the four user groups. User group 4: Fully trained nurses with <10 years of experience, user group 3: nurses with >10 years of experience, user group 3: Physicians in specialty training (e.g., residents), user group 1: Fully trained physicians (e.g., senior physician, board-certified physician, medical specialist, physicians with at least 8 years of working experience)

#### Stethoglove^®^ handling stratified by gender, BMI and auscultated body site of auscultation

The influence of the independent variables gender, BMI, and auscultated organ/body site on the rating of the Stethoglove^®^ handling was evaluated separately. The mean rating of Stethoglove^®^ handling by all users was 3.8 ± 0.9 for males and 3.5 ± 0.7 for females (Median 4.0; range 2-5 for both).

When stratified for BMI, the mean rating of Stethoglove^®^ handling was 3.8 ± 0.8 (for normal weight patients), 3.8 ± 0.9 (for overweight patients), 3.6 ± 0.7 (for class I obesity patients) and 3.2 ± 0.5 (for class III obesity patients).

When stratified for auscultated organ/body site, the mean rating of Stethoglove^®^ handling was 3.7 ± 0.8 for lungs, 3.8 ± 0.8 for hearts, and 3.7 ± 0.8 for the abdomen (Median 4.0; range 2-5 for all). For “other” body sites including the carotid artery, femoral artery and stomach, a lower mean rating was assigned (3.6 ± 0.5), however, the underlying number of auscultations was substantially lower than for the lung, abdomen and heart.

#### Stethoglove^®^ handling by user group

The Stethoglove^®^ handling was rated “reasonable to good” by each of the four user groups (means ranging from 3.3 ± 0.7 to 4.0 ± 0.9).

## DISCUSSION

In this study, the performance and safe use of Stethoglove^®^, a new, non-sterile, single-use hygienic cover for stethoscopes, was demonstrated under real-life conditions in the routine setting of an ICU for postoperative care of cardiac surgery patients. In total, 534 auscultations of different body regions including lungs, heart and abdomen were performed with Stethoglove^®^ and rated for acoustic quality and product handling.

### Stethoglove^®^ offers good acoustic quality of auscultations and an reasonable handling profile

The overall acoustic quality of auscultations with Stethoglove^®^ was “good” as rated by all users with a mean of 4.2 on a 5-point Likert scale (primary performance endpoint). To further explore this result, we evaluated only those auscultations, that were rated “good” (score of 4) or “very good” (score of 5), as these ratings were considered most meaningful in terms of acoustic quality.

A total of 86.1% of all auscultations was rated “good” or “very good” across all users, and this value is in good agreement with the values of each user group (Table 3).

These results are important for different reasons. First, they indicate a high level of satisfaction among all users with regard to the overall quality of auscultation sound transmission through the specific Stethoglove^®^ material. Second, these findings indicate that Stethoglove^®^ is capable of ensuring a high degree of auscultation accuracy in different body regions. This finding is of utmost clinical relevance as data on acoustic quality of commercially available stethoscope covers are scarce. Moreover, other technologies such as fully disposable single-use stethoscopes have been reported to come with substantial impairment of auscultation accuracy which may potentially lead to misdiagnoses and may therefore ultimately even impact patients’ safety ^16,17^.

When stratified for BMI, the ratings ‘good’ or “very good” were recorded for 91.9% of auscultations in normal weight patients. As expected, the rating of the acoustic quality decreased with increasing BMI (85.1% in overweight patients, 85.3% in class I obese patients, and 62.5% in class III obese patients). This observation is not surprising, as with increasing body fat mass in thoracic or abdominal body areas, heart, lung or gastrointestinal tones would appear muted during normal auscultations. This observation is thus not attributable to the medical device.

When stratified for gender, the percentage of all auscultations rated as “good” and “very good” was 81.6% in females and 87.7% in males. Both values are in good agreement with the overall rating by all users (Table 3). The slight difference between males and females might be rather due to their anatomical differences in the upper thoracic area, rather than device-related.

When stratified for auscultated body site the rating of the acoustic quality was stated as “good” and “very good” for 87.6%, 89.2%, and 84.2% of all auscultations of the heart, lung, and abdomen. These results are in good agreement with the overall rating by all users (Table 3). Very few auscultations (N=10) were performed on “Other” body sites including carotid and femoral arteries and stomach. The percentage of auscultations rated as “good” or better was lower (40.0%) than for heart, lung, or abdomen. However, this finding is not unexpected and not considered to be attributable to Stethoglove^®^ given the general challenge in auscultating these “Other” body sites and their generally low sound profile, especially if no pathological alterations have occurred.

In regard to the secondary performance endpoint, this study showed that Stethoglove^®^ is easy to handle and may thus enable quick implementation into the clinical routine of HCPs. Handling of the product was rated for 96.4% of all auscultations to be “reasonable”, “good” or “very good” (score of 3, 4 or 5 on the 5-point Likert scale), highlighting the high degree of acceptance by the users and their general willingness to use Stethoglove^®^ in their daily work.

The stratification analysis showed that despite some expected differences between males and females, body sites as well as among BMI classes, which do all apply to auscultations in general, Stethoglove^®^ presented with an overall good performance profile throughout the study and for all patients independent from gender, BMI or auscultated body site. Even in highly obese patients, for which it is well known that auscultations can be truly challenging, the device performed in a satisfying manner based on the user ratings.

### Stethoglove^®^ can be quickly adopted into clinical routine

The high level of acceptance for the product observed in the present study is encouraging when considering the generally extremely low compliance of HCPs towards sufficient stethoscope hygiene or the general reluctance to adopt new technologies or procedures to improve it. Even though stethoscope cleanliness guidelines exist ^13^, and despite tremendous educational efforts, this problem remains a key challenge. In fact, most of the approaches taken to enhance stethoscope hygiene have been ineffective and ultimately failed ^2,11,12^. In a recent observational investigation, Boulee et al. evaluated physicians’ frequency and methods of stethoscope and hand hygiene practices. In only 18% of the 400 observed interactions, stethoscopes were cleaned at all, and when done, less than 4% complied with the CDC guidelines ^13^. The authors concluded that stethoscope hygiene is largely neglected in daily clinical routine and they strongly urged that the compliance towards stethoscope cleanliness needs to be rapidly addressed ^11^. To this end, Muniz and colleagues assessed the reasons for poor stethoscope hygiene compliance and anonymously surveyed nurses, nurse practitioners, and physicians at a large academic pediatric hospital. They found that perceived barriers for low stethoscope hygiene included lack of materials on hand, lack of time, and most importantly, lack of visual reminders^12^. Hence, the high level of acceptance of Stethoglove^®^ seen in this study provides encouraging evidence that the Stethoglove^®^ product may be quickly and efficiently adopted by HCPs to become part of their clinical routine in order to help improving patient protection from stethoscope-mediated infections.

Another interesting observation from this study was the rapid learning curve of the Stethoglove^®^ application which substantially differs from other stethoscope single-cover concepts that usually target the sole coverage of the stethoscope diaphragm. Stethoglove^®^, however, covers the stethoscope bell including the users auscultating hand. After its use, it is easily pulled off the hand (which is still holding the stethoscope) and discarded. In fact, the Stethoglove^®^ concept is based on the same principles as other single-use protective equipment that is used in clinical routine (e.g., medical gloves). Notably, this may represent a conceptual advantage when compared to other commercially available standard microbiological stethoscope barriers as these require manual placement of the clean cover on a potentially contaminated stethoscope. These maneuvers carry a considerable risk for iatrogenic contamination of either the clean disc, the user’s hand, or both. To the contrary, this risk could be minimized or even fully eliminated with the Stethoglove^®^ approach.

## LIMITATIONS

This study has several limitations: Firstly, this was a single-center study in an ICU setting and user experiences with the product in other healthcare settings are not available yet.

However, this study was carried out in a high-demand ICU environment on early postoperative cardiac patients, i.e., a critical patient population. Therefore, although results cannot be generalized to other healthcare settings and patient populations, expansion of observations

from this study to other health care areas is anticipated. Secondly, the evaluation of prevention of pathogen transmission was not assessed, and was beyond the scope of this study. Thirdly, to enable a real-life setting, no control group or randomization was added to the study protocol. In addition, auscultating the same patient with and without Stethoglove^®^ even in a randomized manner would have caused a bias as the user would have known what to hear after the first auscultation event (whether with or without the Stethoglove^®^ cover).

Finally, it should also be recognized that the study was carried out on postoperative cardiac patients in a high demand ICU environment. For such freshly operated patients, it is well established that the proper auscultation can be challenging. They are often lying flat in their ICU beds, and usually still carry loads of fluids in their body (e.g., in the pleura or pericardium or abdominally) early after surgery, thus representing a “worst case scenario” population for auscultation.

## CONCLUSION

This study using a real-world healthcare environment of a cardiovascular ICU demonstrated that Stethoglove^®^ can be safely and effectively used as a hygienic cover for stethoscopes during auscultations. Device acceptance was high across various HCP user groups indicating the device’s potential of being quickly adopted into daily clinical routine. The rating levels for the acoustic quality of auscultations performed with Stethoglove^®^ were high across all user groups. Effects of gender, auscultation site and BMI on the acoustic quality of auscultations Stethoglove^®^ were comparable to the effects on auscultations in general, but not attributable to the use or design of the product. Therefore, Stethoglove^®^ may represent a safe, simple and useful tool to improve patient safety and to reduce the risk of stethoscope-mediated infections in the clinical routine workflow without impairing the acoustic quality of auscultations.

## Data Availability

All data produced in the present study are available upon reasonable request to the authors

## ABBREVIATIONS

BMI: Body-Mass-Index
HCI: Healthcare-associated infection
HCP: Healthcare professional
ICU: Intensive care unit
TPU: Thermoplastic urethane

## ACKNOWLEDGEMENT

The authors of this manuscript would like to thank the nursing and physician staff of the intensive care unit at the German Heart Center Berlin for their active and voluntary participation in this study. TNS is a scholar in the BIH Charité Clinician Scientist Program funded by the Charité–Universitátsmedizin Berlin and the Berlin Institute of Health.

## AUTHOR CONTRIBUTION STATEMENT

TNS and MYE designed and oversaw the clinical study and prepared the first draft of this manuscript. HM, VE, MS, FS, AF and TNS were responsible for patient recruitment and data management. All authors contributed and approved the final draft of this manuscript. Data analysis was performed by Heidi Kulas from the IGES Institute GmbH in Berlin.

## DATA AVAILABILITY STATEMENT

Data are made available upon reasonable request.

